# Excessive daytime sleepiness among dental students: A meta-analysis of prevalence

**DOI:** 10.1101/2023.12.19.23300251

**Authors:** Jorge Homero Wilches-Visbal, Alex Antonio Angulo-Luna, Adalberto Campo-Arias

## Abstract

**Introduction:** Excessive daytime sleepiness is highly prevalent and variable in health students. However, a meta-analysis that synthesizes the prevalence of this symptom in dental students has yet to be reported.

**Objective:** To compute the pooled prevalence of sleepiness in dental students.

**Method:** A systematic review and meta-analysis of studies published between 2000 and 2023 was developed in Spanish, English, and Portuguese, with the keywords sleepiness, excessive drowsiness, Epworth scale for drowsiness, and dental students in all three languages. Original articles and degree theses that reported the frequency of sleepiness with a cut-off point for the Epworth scale for sleepiness greater than ten were included. The sample sizes and the number of students positive for sleepiness were observed, and the prevalence, raw and adjusted for sample size, was calculated.

**Results:** Seven articles were included with participant samples between 128 and 325 students, totaling 1,546. Students were positive on the Epworth scale for sleepiness between 43 and 189, with a sum of 671. The pooled prevalence was 42.7% (95% CI 40.3-45.2).

**Conclusions:** drowsiness is present in four out of ten dental students. It is necessary to systematize the factors associated with drowsiness and establish a relationship with the academic performance of dental students.

## INTRODUCTION

Drowsiness is the propensity to sleep during waking hours.^1,2^ Excessive daytime sleepiness frequently occurs in idiopathic hypersomnolence, narcolepsy, Kleine-Levin syndrome, and obstructive sleep apnea/hypopnea.^3,4^ Likewise, people who meet the criteria for mental disorders such as anxiety disorders, depressive disorders, and primary insomnia often complain of excessive sleepiness.^1,5-7^

Short-term sleepiness is associated with a deterioration in cognitive performance that affects attention, memory, and learning functions and, consequently, can affect students’ academic performance.^8,9^ Furthermore, sleepiness can cause mood alterations, induce the abuse of stimulant substances, and predispose to traffic accidents.^10,11^ In the long term, sleepiness increases the risk of cardiovascular disease and obesity, among others.^12-14^

On the other hand, sleepiness in adolescent and young adult students, in most cases, is due to poor sleep hygiene secondary to behavioral changes to meet schedules and other academic demands.^8-11,15^

In medical students, two systematic reviews showed a high prevalence of sleepiness by applying the Epworth scale for sleepiness. Jahrami et al.^16^ included nine studies that included 2,587 participants and found a pooled prevalence of sleepiness of 34.6% (95% CI 18.3-50.9). More recently, Binjabr et al.^17^ reviewed 28 studies involving 10,122 students and reported a pooled prevalence of sleepiness of 33.3% (95% CI 26.5-40.9). However, it is necessary to have a systematic review of the prevalence of drowsiness in dental students. Knowing an accurate and reliable estimate of the prevalence of sleepiness is vital to identifying possible associated risk factors and developing prevention, early identification, and management strategies for sleepiness specific to dental students’ context and particular needs.^4,18-20^

This systematic review and meta-analysis aimed to estimate the pooled prevalence of sleepiness among dental students.

## METHODS

A systematic review and meta-analysis of primary studies reporting the prevalence of sleepiness in dental students were conducted.

### Search for relevant studies

A scientific literature search was carried out using specific descriptors in Spanish, English, and Portuguese combined using the Boolean operators ‘AND’, and ‘OR’. The search was carried out in Google Scholar and PubMed, which bring together the most significant number of relevant sources for studies in the health area. Equivalents to “sleepiness” and “Epworth” were used as descriptors in the languages as mentioned earlier. “Sleepiness scale,” “dentistry students,” “odontology students,” and “dental students.” The search equation used was: “Epworth sleepiness scale” OR “sleepiness” AND “dentistry students” OR “odontology students” OR “dental students”.

### Inclusion and exclusion criteria

The authors included original articles and graduate papers published from January 2000 to October 2023 that used the Epworth Sleepiness Scale. Those studies were excluded in which the measurement of sleepiness omitted information on the prevalence of sleepiness, or the data presented prevented the calculation of precise data on participants with excessive daytime sleepiness with a cut-off point greater than ten.^21,22^

Narrative reviews, citations, and results from gray literature, such as reports, documents, or manuals usually published without peer review, were excluded.^23^

### Data extraction

Initially, the titles of the studies were examined. Subsequently, two authors independently reviewed the abstracts, and articles were selected after reading the methods and results sections. Finally, the sample sizes the number of students positive for drowsiness, were observed.

The quality of the studies was quantified with a critical appraisal instrument for scientific reports.^24^ This measurement instrument summarizes the STROBE criteria for observational studies.^25^

### Heterogeneity assessment and bias control

A requirement for conducting a meta-analysis is for the group of included studies to be sufficiently homogeneous regarding participants and results. The variability between studies is heterogeneous and may be due to methodological diversity. Heterogeneity (tau) was assessed using I;^2^ values greater than 50% are considered high.^26^

Publication bias was assessed using the Rosenthal fail-safe N method.^27^ This method identifies the number of negative studies (with zero effect) that would need to be added for the meta-analytic result to be null. If the p-value associated with the fail-safe N value is more significant than 0.05, it is considered that there is publication bias.^27^ Despite being the most used parameter to evaluate publication bias, fail-safe N is highly variable with the method.^28^ This test assumes that all non-significant effect sizes are equal to zero, regardless of sample size, and focuses more on statistical significance than on effect magnitude.^29^

### Data synthesis

The pooled prevalence of sleepiness was calculated with 95% confidence intervals (95%CI) with the application of the inverse variance method.^30^ When heterogeneity is low, applying the fixed effects model is suggested. On the contrary, the random effects model is indicated if heterogeneity is high.^31^ The analysis was completed with the free program Jamovi version 2.3.^32^

## RESULTS

One hundred and fifty-three titles were identified, of which ten were duplicated in Google Scholar. Only one study was identified in Pubmed, which was already identified in Google Scholar. After reading the titles and abstracts, thirteen articles were selected, whose full text was reviewed. Of these, six articles were ignored because they omitted information on the prevalence of sleepiness, and it was impossible to calculate it with the information presented.^33-38^ No additional titles were identified in the articles cited in the references of the initially included studies. Consequently, seven that met the inclusion criteria were reviewed.^39-45^ The complete flowchart of the selection process is presented in Figure 1. The summary of the seven cross-sectional analyses that reported the prevalence of sleepiness in dental students is presented in Table 1.

**Table 1.** Summary of the included papers.

| Authors | Country | Sample | Age*<br>Range | Mean | Women<br>n (%) | Epworth<br>>10<br>n (%) |
| --- | --- | --- | --- | --- | --- | --- |
| Figueroa et al.<sup>39</sup> | Chili | 225 | 18-28 | 23.1 | 149 (66.2) | 93 (41.3) |
| Angelin et al.<sup>40</sup> | Brazil | 325 | 18-37 | NR | 251 (77.2) | 189 (58.2) |
| Gonçalves et al.<sup>41</sup> | Brazil | 221 | NR | NR | NR | 129 (58.4) |
|  | Brazil | 136 | NR | 21.3 | 101 (74.3) | 56 (41.2) |
| Santos et al.<sup>42</sup> | Malaysia | 198 | 20-26 | NR | 146 (73.7) | 90 (45.5) |
| Ramli et al.<sup>43</sup> | Brazil | 128 | <18 | 21.0 | 97 (75.8) | 43 (33.6) |
| Turcio et al.<sup>44</sup> | Türkiye | 313 | 18-40 | 21.8 | 195 (62.3) | 71 (22.7) |
| Gaş et al.<sup>45</sup> |  |  |  |  |  |  |

\*Expressed in years. NR did not report. None of the studies reported the Epworth Sleepiness Scale's internal consistency (Cronbach's alpha).

**Figure 1.**
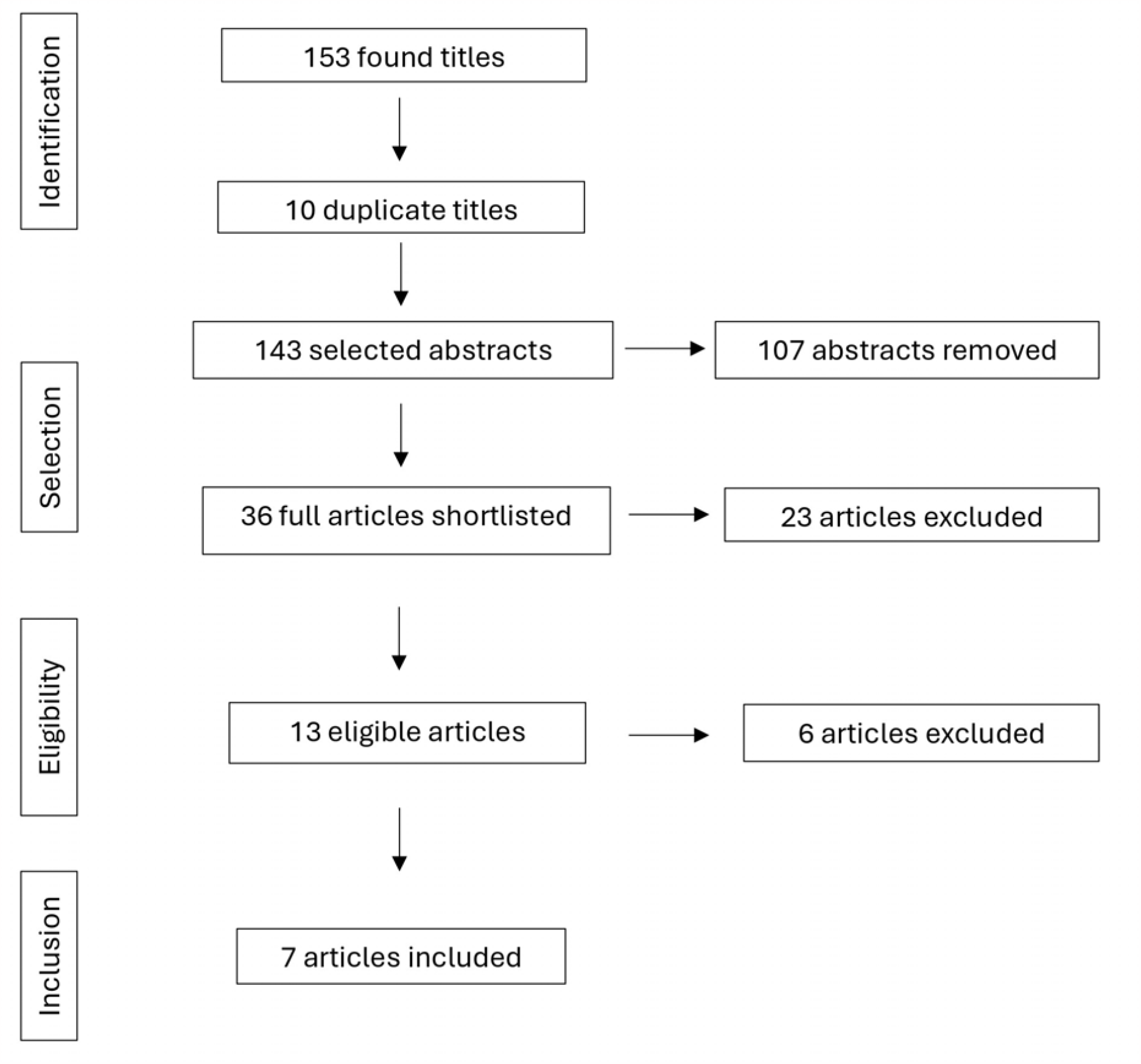
Flowchart of the articles.

The sample size of participants for the seven included papers varied between 128 and 325 students, with a total of 1,546 students. The positive students on the Epworth scale for sleepiness were between 43 and 189, totaling 671 students.

The age of the participants varied between 18 and 40 years; only one study included participants under 18 years of age.^44^ Between 62 and 77% of the students positive for the Epworth scale were female students.

The studies showed high heterogeneity, with a tau value of 0.13 and I^2^ of 93.8% (p<0.001). This finding was corroborated in the funnel plot (see graph 2), and the fail-safe N value was 3,100 (p<0.01). Therefore, the pooled prevalence was calculated with the random method and showed a value of 42.7% (95% CI 40.3-45.2). See details in graph 3.

**Graph 2.**
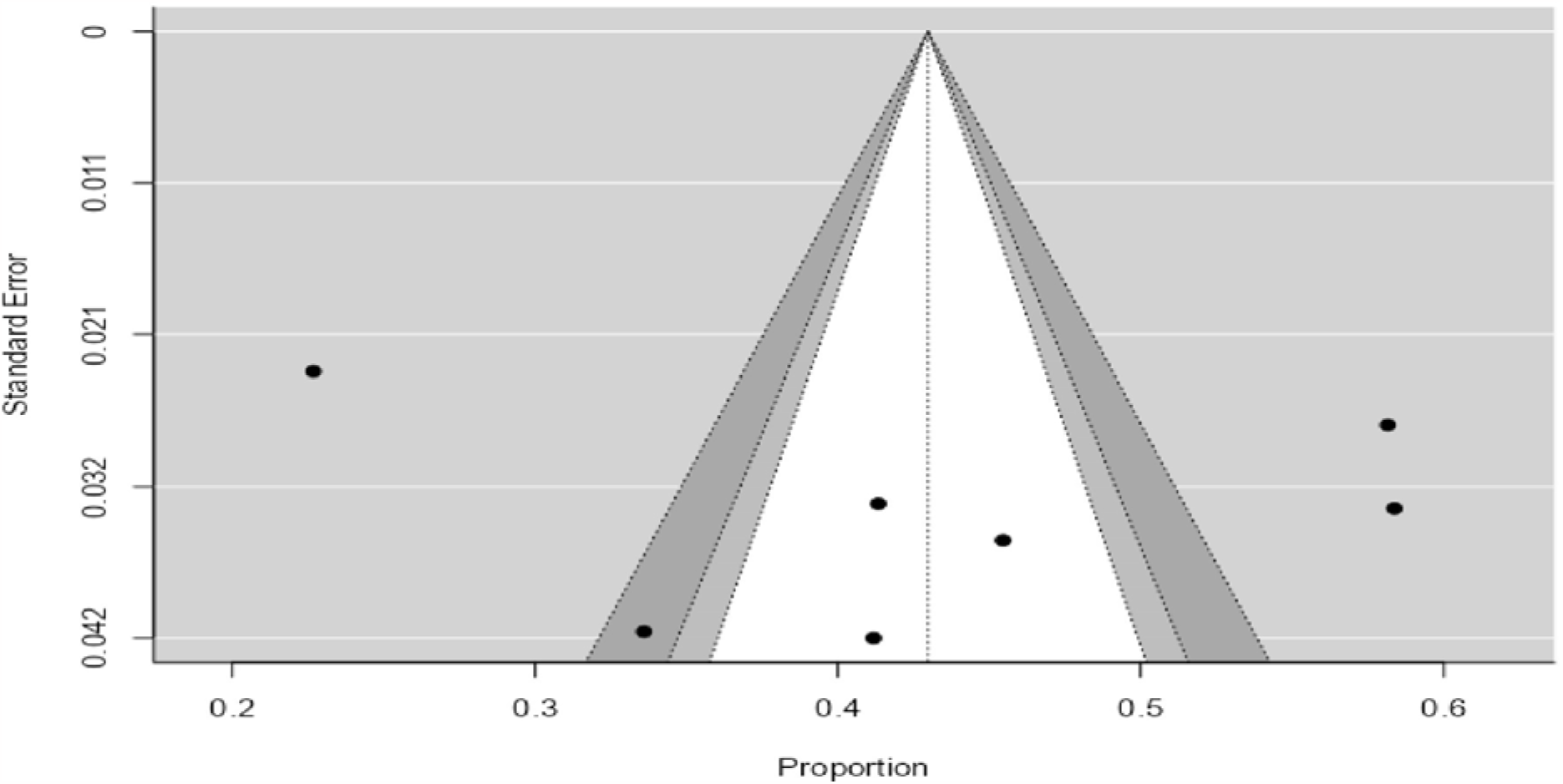
Funnel plot for heterogeneity.

**Graph 3.**
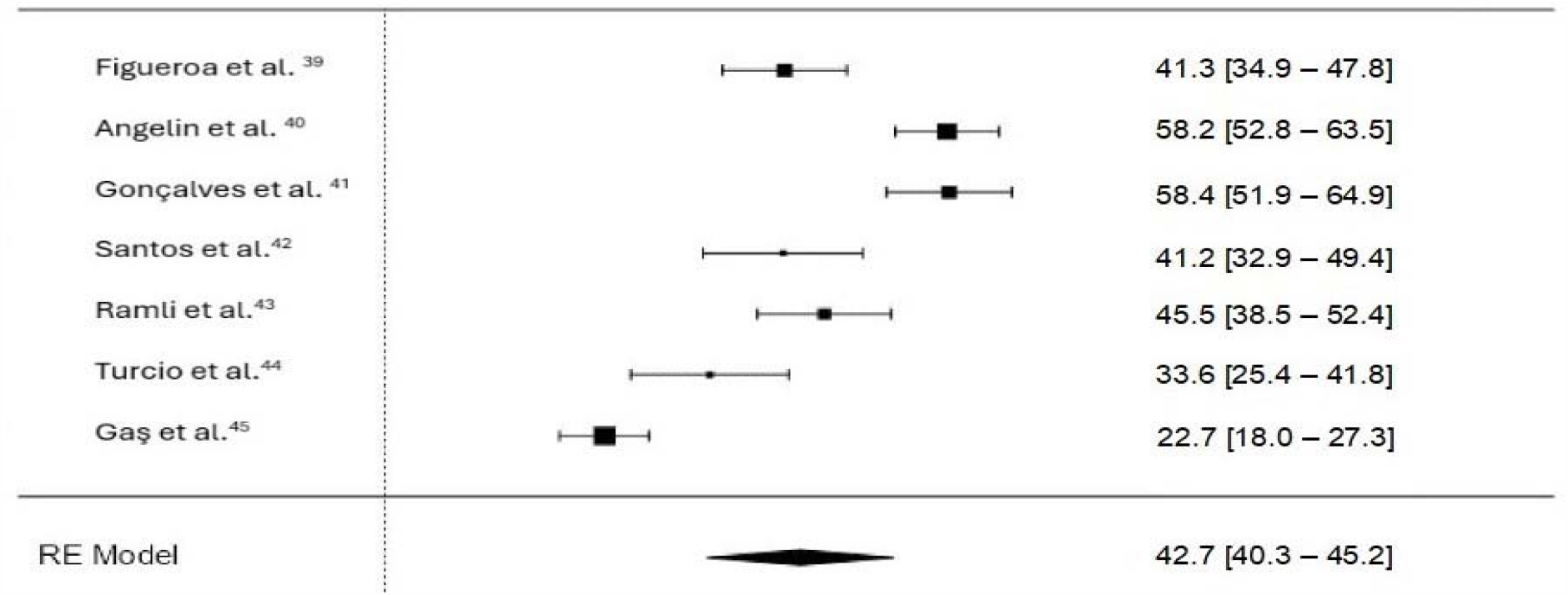
Forest plot for the prevalence of sleepiness.

## DISCUSSION

The present review shows that approximately four in ten dental students report drowsiness. This grouped prevalence was higher than that observed in other meta-analyses of sleepiness in medical students, in which prevalences of around 43% were observed.^16,17^

The high prevalence of drowsiness in dental students is a multifactorial problem, as in other university students. ^1^ Sleepiness in this particular population can be explained by a considerable deterioration in sleep hygiene with entry to higher education, such as a decrease in sleeping hours or irregular sleep patterns due to early classes, university social lifestyle, and academic demands.^8-11,15^ To this can be added other psychosocial stressors that increase the risk of anxiety, depression, and insomnia that usually occur with drowsiness.^1,5-7^ It is necessary to keep in mind that sleepiness can be a symptom of mental disorders such as anxiety and depression, which are highly frequent in university students in the global context.^46-50^ Therefore, sleepiness among university students requires a careful evaluation of all possible causes to define the best approach. ^51^

In general, university students report a higher prevalence of sleepiness than the general adult population, in whom the frequency of sleepiness can reach 12%.^52^ University students should be considered a vulnerable population at high risk of drowsiness, and, consequently, university authorities should implement screening strategies in the university context^18-20^ and activate the care route to identify the causes and provide the best solutions and treatment options for everyone.^51^ These early actions to recognize these cases can mitigate the adverse effects of drowsiness on academic performance and future professional practice.^8,9^

This review has the advantage of including works published in Spanish and Portuguese that should be addressed in reviews carried out by English-speaking authors. Furthermore, the review covered gray literature, which reduces publication bias.^53^ However, this meta-analysis had the limitation that the included studies should have included more information on the validity and reliability of the Epworth scale for sleepiness in the participating samples. This information is relevant because the reliability indicators of measurement scales can vary dramatically between populations and compromise the internal validity of the findings.^54^ Likewise, the high heterogeneity of the research could be considered a limitation. However, calculating the grouped prevalence by the random method makes an essential adjustment to this indicator.^31^

It is concluded that drowsiness affects 42.7% of dental students. It is necessary to systematize the factors associated with drowsiness and establish a relationship with the academic performance of dental students. It is mandatory to manage strategies for early identification in the university context and refer for formal diagnosis and timely management.

## Data Availability

The data supporting this study's findings are available from the corresponding author upon reasonable request.

## Notes

**Conflict of interest:** The authors declare no interest in the research.

**Data availability statement:** The data supporting this study’s findings are available from the corresponding author upon reasonable request.

### Competing Interest Statement

The authors have declared no competing interest.

### Funding Statement

This study was funded by the Universidad del Magdalena, Santa Marta, Colombia.

## REFERENCES

1. Gandhi KD, Mansukhani MP, Silber MH, Kolla BP. Excessive daytime sleepiness: A clinical review. Mayo Clin Proc. 2021;96(5):1288–301.

2. Martin VP, Lopez R, Dauvilliers Y, Rouas JL, Philip P, Micoulaud-Franchi JA. Sleepiness in adults: An umbrella review of a complex construct. Sleep Med Rev. 2023;67:101718.

3. He K, Kapur VK. Sleep-disordered breathing and excessive daytime sleepiness. Sleep Med Clin. 2017;12(3):369–82.

4. Hartman L, Hook W. Sleep disorders & treatment. Osteopat Fam Phys. 2017;9(3):30–8.

5. Simon EK, Berki ZH, Gettys GC, Vedak C. Sleep problems and disorders in patients with anxiety disorders. Psychiatr Ann. 2016;46(7):396–400.

6. Hein M, Lanquart JP, Loas G, Hubain P, Linkowski P. Prevalence and risk factors of excessive daytime sleepiness in insomnia sufferers: A study with 1311 individuals. J Psychosom Res. 2017;103:63–9.

7. Hein M, Lanquart JP, Loas G, Hubain P, Linkowski P. Prevalence and risk factors of excessive daytime sleepiness in major depression: A study with 703 individuals referred for polysomnography. J Affect Disord. 2019;243:23–32.

8. Hershner SD, Chervin RD. Causes and consequences of sleepiness among college students. Nature Sci Sleep. 2014;6:73–84.

9. Suardiaz-Muro M, Morante-Ruiz M, Ortega-Moreno M, Ruiz MA, Martín-Plasencia P, Vela-Bueno A. Sueño y rendimiento académico en estudiantes universitarios: revisión sistemática. Rev Neurol. 2020;71:43–73.

10. Moore M, Meltzer LJ. The sleepy adolescent: Causes and consequences of sleepiness in teens. Paediatr Respir Rev. 2008;9(2):114–21.

11. Merdad RA, Akil H, Wali SO. Sleepiness in adolescents. Sleep Med Clin. 2017;12(3):415–28.

12. Mustač F, Matovinović M, Mutak T, Barun B, Jug J, Levicki R, et al. Excessive daytime sleepiness as cardiovascular risk in Croatian obese patients. Cardiol Croat. 2019;14(9-10):236.

13. Wang L, Liu Q, Heizhati M, Yao X, Luo Q, Li N. Association between excessive daytime sleepiness and risk of cardiovascular disease and all-cause mortality: A systematic review and meta-analysis of longitudinal cohort studies. J Am Med Direct Assoc. 2020;21(12):1979–85.

14. Li J, Covassin N, Bock JM, Mohamed EA, Pappoppula LP, Shafi C, et al. Excessive daytime sleepiness and cardiovascular mortality in US adults: A NHANES 2005– 2008 follow-up study. Nature Sci Sleep. 2021;13:1049–59.

15. Yeo SC, Lai CK, Tan J, Lim S, Chandramoghan Y, Tan TK, et al. Early morning university classes are associated with impaired sleep and academic performance. Nature Hum Behav. 2023;7(4):502–14.

16. Jahrami H, Alshomili H, Almannai N, Althani N, Aloffi A, Algahtani H, et al. Predictors of excessive daytime sleepiness in medical students: A meta-regression. Clock Sleep. 2019;1(2):209–19.

17. Binjabr MA, Alalawi IS, Alzahrani RA, Albalawi OS, Hamzah RH, Ibrahim YS, et al. The worldwide prevalence of sleep problems among medical students by problem, country, and COVID-19 status: A systematic review, meta-analysis, and meta-regression of 109 studies involving 59427 participants. Curr Sleep Med Report. 2023;9:161–79.

18. Shepardson RL, Funderburk JS. Implementation of universal behavioral health screening in a university health setting. J Clin Psychol Med Set. 2014;21:253–66.

19. Friedrich A, Schlarb AA. Let’s talk about sleep: A systematic review of psychological interventions to improve sleep in college students. J Sleep Res. 2018;27(1):4–22.

20. Prichard JR, Hartmann ME. Follow-up to Hartmann & Prichard: Should universities invest in promoting healthy sleep? A question of academic and economic significance. Sleep Health. 2019;5(4):320–5.

21. Johns MW. A new method for measuring daytime sleepiness: The Epworth sleepiness scale. Sleep. 1991;14(6):540–5.

22. Johns MW. Reliability and factor analysis of the Epworth Sleepiness Scale. Sleep. 1992;15(4):376–81.

23. Montes de Oca JL. La literatura gris cambia de color: un enfoque desde los problemas sociales de la ciencia y la tecnología. MediSur. 2018;16(3):424–36.

24. Prel JB du, Röhrig B, Blettner M. Critical Appraisal of Scientific Articles. Dtsch Arztebl Int. 2009;106(7):100–5.

25. Von Elm E, Altman DG, Egger M, Pocock SJ, Gøtzsche PC, Vandenbroucke JP. The Strengthening the Reporting of Observational Studies in Epidemiology (STROBE) Statement: Guidelines for Reporting Observational Studies. Ann Intern Med. 2007;147(8):573–78.

26. Fletcher J. What is heterogeneity and is it important? BMJ. 2007;334(7584):94–6.

27. Rosenthal R. The ‘‘file drawer problem’’ and tolerance for null results. Psychol Bull. 1979;86:638–41.

28. Rosenberg MS. The file-drawer problem revisited: A general weighted method for calculating fail-safe numbers in meta-analysis. Evolution 2005;59(2):464–8.

29. Van Aert RCM, Wicherts JM, van Assen MA. Publication bias examined in meta-analyses from psychology and medicine: A meta-meta-analysis. PLoS One. 2019;14(4):1–32.

30. Barendregt JJ, Doi SA, Lee YY, Norman RE, Vos T. Meta-analysis of prevalence. J Epidemiol Community Health. 2013;67(11): 974–8.

31. Christoffersen MN, Poulsen HD, Nielsen A. Attempted suicide among young people: risk factors in a prospective register based study of Danish children born in 1966. Acta Psychiatr Scand. 2003;108(5):350–8.

32. The jamovi project. Jamovi (Version 2.3); 2022. Retrieved from https://www.jamovi.org.

33. Coutinho ALL, Pinto G. A influência da época de exames na ansiedade e na qualidade do sono dos alunos da Faculdade de Medicina Dentária da Universidade de Lisboa (Doctoral dissertation). Lisboa: Universidade de Lisboa; 2019.

34. Avanak SN, Avanaki NN, Soleimani P, Rafiei H. Prevalence of daytime sleepiness among medical university students. J Prev Epidemiol. 2018;3(2):e09.

35. Nurismadiana I, Lee K. Factors associated with sleep quality among undergraduate students at a Malaysian public university. Int J Public Health Clin Sci. 2018;5(6):373.

36. Dias FI, Cardoso B, de Souza L, Batista E, Pigossi SC, Santana L. Perception of health-related quality of life, sleep quality and sleepiness index in an educational environment at a dental school in Southeast Brazil. Eur J Dent Educ. 2022;26(4):794–800.

37. Dias FI, Silva IC, Trubucci J, da Silva KC, Magalhães AL, Alvitos R, Santana L. Evaluation of quality of life, sleep, and sleepiness in dental students during active learning and remote emergency learning. Rev Cienc Saude. 2023;13(3):17–22.

38. Norman P, Karthikeyan E, Thirunaaukarasu D, Hafeez S. Sleep deprivation and its association with depression in first-year medical and dental students in Kanchipuram District, Tamil Nadu, India. Orapuh J. 2023;4(1):e1004.

39. Figueroa BA, González Á. Relación de la percepción de calidad de sueño y dolor orofacial en estudiantes de Odontología. Estudio de cohorte prospectivo (Doctoral dissertation). Talca: Universidad de Talca; 2018.

40. Angelin TJ, Rodrigues K, Dos Santos VE Jr., Cosme L, Vilela M. Evaluation of sleep quality and daytime sleepiness in dentistry students. Pesq Bras Odontopediatr Clínica Integr. 2020;20:e0003.

41. Gonçalves RA, Reis M, Trento G, Spin-Neto R, Marques M, Pereira-Filho VA. Evaluation of daytime sleepiness and academic performance in dentistry students. Eur J Gen Dent. 2021;10(01):37–43.

42. Santos A, Esteve N, Matos E, Bassi D, Resende V, de Jesus RR, et al. O impacto do Covid-19 na qualidade do sono, grau de estresse e rotina de estudo de acadêmicos de odontologia. Res Soc Dev. 2021;10(6):e51910616073.

43. Ramli R, Hasbullah N, Ghani N. Excessive daytime sleepiness and academic performance among dental students in north-east of peninsular Malaysia. Asian J Med Biomed. 2022;6(1):48–56.

44. Turcio KH, de Moraes-Melo-Neto CL, de Caxias FP, Goiato MC, Dos Santos DM., Januzzi MS et al. Relationship of excessive daytime sleepiness with bruxism, depression, anxiety, stress, and sex in odontology students–A cross sectional study. J Clin Exp Dent. 2022;14(6):e464.

45. Gaş S, Yildirim, G. Assessment of the relationship between obstructive sleep apnea syndrome and sleep quality among dental students. J Health Sci Med. 2023;6(5):981–6.

46. Sarokhani D, Delpisheh A, Veisani Y, Sarokhani MT, Manesh RE, Sayehmiri K. Prevalence of depression among university students: A systematic review and meta-analysis study. Depress Res Treat. 2013;2013:373857.

47. Shaffique S, Farooq SS, Anwar H, Asif HM, Akram M, Jung SK. Meta-analysis of prevalence of depression, anxiety and stress among university students. RADS J Biol Res Appl Sci. 2020;11(1):27–32.

48. Akhtar P, Ma L, Waqas A, Naveed S, Li Y, Rahman A, et al. Prevalence of depression among university students in low and middle income countries (LMICs): A systematic review and meta-analysis. J Affect Disord. 2020;274:911–9.

49. Gao L, Xie Y, Jia C, Wang W Prevalence of depression among Chinese university students: A systematic review and meta-analysis. Sci Rep. 2020;10(1):15897.

50. Ahmed I, Hazell CM, Edwards B, Glazebrook C, Davies EB. A systematic review and meta-analysis of studies exploring prevalence of non-specific anxiety in undergraduate university students. BMC Psychiatry. 2023;23(1):340.

51. Peter-Derex L, Micoulaud-Franchi JA, Lopez R, Barateau L. Evaluation of hypersomnolence: From symptoms to diagnosis, a multidimensional approach. Rev Neurol (Paris). 2023;179:715–26.

52. Ford ES, Cunningham TJ, Giles WH, Croft JB. Trends in insomnia and excessive daytime sleepiness among US adults from 2002 to 2012. Sleep Med. 2015;16(3):372–8.

53. Paez A. Gray literature: An important resource in systematic reviews. J Evidence-Based Med. 2017;10(3):233–40.

54. Cohen L, Manion L, Morrison K. Validity and reliability. In: Cohen L, Manion L, Morrison K. Research methods in education. 8th edition. London: Routledge; pp. 245–84.

